# Dataset on the COVID-19 Pandemic Situation in Tunisia with Application to SIR Model

**DOI:** 10.1101/2020.04.23.20076802

**Authors:** Abir Lassoued, Afef Ben Saad, Hela Lassoued, Raouf Ketata, Olfa Boubaker

**Author notes:** **Corresponding author(s)** Olfa Boubaker.

## Abstract

April 9, 2020 marks 100 days since the first cases of coronavirus disease 2019 in China. In this crucial day with 1 436 198 confirmed infected cases in the world and 85 521 deaths, the Global Level of the Covid-19 pandemic was evaluated at very high according to the World Health Organization (WHO) situation report. For most people, COVID-19 infection will cause mild illness (fever and at least one symptom of respiratory disease); however, for more than 3.4% of people, it can be fatal. Older people and those with preexisting medical conditions (such as cardiovascular disease, chronic respiratory disease, or diabetes) are at risk of severe disease and mortality. The incubation period of the virus is estimated to be between 2 and 14 days, but longer incubation had been reported. Furthermore, data published by world authorities show that statistics are different for different geographical regions and depend on many social and environmental factors. The sad reality of the COVID-19 is that there are currently no medications or vaccines proven to be effective for the treatment or prevention of the disease. The pandemic spread is consequenctly followed by a worldwide panic. Facing this dramatic uncertain situation, implementing a country-wide strategy for social distancing and a general logistic policy for critical and life-saving supplies is an urgent for government and sanitary authorities. Several mathematical models have been proposed to predict epidemic spread. However, models should be adapted to specific situations in countries where geographic, societal, economic, and political strategies are different. Here, we propose the application of the well-known SIR model to the case study of Tunisia for which data are collected from three databases in order to have rapidly predict the situation. Such results can be useful in the future to design a more reliable model to help in monitoring infection control.

## Data accessibility

Data can be retrieved from the public Data repository Mendeley at https://data.mendeley.com/datasets/v8fhn6wsh2/3 [1]. Data were collected from three sources [2–4] and then compiled using Excel software and the open source MATLAB code available at [5].

## Value of the Data

- These data are useful because they represent a simple approach to evaluate the transmission dynamics of COVID-2019 in developing countries, for which very few results are reported.
- Governmental authorities as well public health institutes can benefit from these data since they can predict the epidemic spread and implement a country-wide strategy for social distancing and logistic policy of critical and life-saving supplies. Such tools can also be used for medical education and biomedical engineering training.
- The additional value of these data lies in the fact to estimating the country unknown parameters related to the SIR model because these parameters are special for different geographical regions and depend on many social and environmental factors.
- The tool can be very useful in designing key strategies to tackle the spread. It can help to verify whether the extensive measures imposed by authorities like social distancing, quarantine, lockdown, and curfew and so on are proficient. The tool also enables the projecting of essential supply needs, including the estimation of biomedical equipment, drugs for supportive care, and consumable medical supplies.
- The collected and compiled data can be used to design a more reliable model including every societal facts and the entire measures taken by authorities in order to estimate the finale size of the spread and propose an objective tool for monitoring infection control. All these facts and measures can be found in [6].

## Experimental Design and Methods

The data of COVID-19 in Tunisia from March 02, 2020 to April 19, 2020 were collected from the website of three official sources: the National Observatory of New and Emerging Diseases of the Tunisian Ministry of Health [3], the Coronavirus Worldometer [4], and the World Health Organization (WHO) [2]. An Excel file was used to build a time-series database [1] including for each source the time series related to the total infected cases, the infected cases/day, the total deaths, and the new daily deaths/day. The daily tests and the total recovered are only collected from the country official source. The well-known Susceptible-Infectious-Recovered (SIR) model is considered in this study to predict the epidemic dynamics in the country [8].

The SIR model is a compartmental model described by three ordinary differential equations that can estimate the number of susceptible (S), the number of infectious (I), and the number of recovered patients (R) in epidemic situations. It is generally used to characterize early spread growth [7,8]. The model is characterized by four parameters to be esteemed: the transmission rate, the average recovery rate, the population size N, and the initial number of infected cases. The model is considered under a number of assumptions; N is constant and does not represent the population size. It is estimated as N= S + I + R. The initial recovered population is assumed zero, and the final number of infected people is also assumed to be zero.

For the implementation of the SIR model in the case study of Tunisia, the Excel file building the database [1] and the open-source MATLAB code [5] are used in order to provide graphical visualizations and short-term forecasts. First of all, the collected data were compiled using the open source code. Then, estimation of the parameters and initial values of the SIR model are obtained by minimizing the differences between the actual and predicted numbers of infected cases. The optimization toolbox via the fminsearch function is used to compute the optimal values of unknown model parameters related to the country. The SIR differential system is finally solved using the MATLAB function *ode45*.

## Results and Analysis

In this paper, the time series of the total infected cases are shown in Fig. 1 and exhibit differences in reported data for the three sources [2–4]. Indeed, data are analyzed via statistical functions and show different standard deviations for the three sources [1]. These differences are essentially due to differences in reporting methods, retrospective data consolidation, and reporting delays. Fig. 2. shows the time series of the total infected cases, the total deaths, and the total recovered collected from the country official source, whereas Fig. 3 shows the time series of the daily tests and the infected cases/day. This later can be helpful in the future to analyze and evaluate the test strategy. To understand the magnitude of the risk posed by this novel coronavirus for the country at this date, the transmission rate (R_0_) computed at the date of April 19, 2020, and represents the number of newly infected people from a single case and the fatality rate (CFR) computed for the same date are found [1]:

*R*_0_ (%) = (total cases/total tests) ×100= 5.11 %
*CFR* (%) = (total death /total cases) ×100= 4.29%

**Figure 1.**
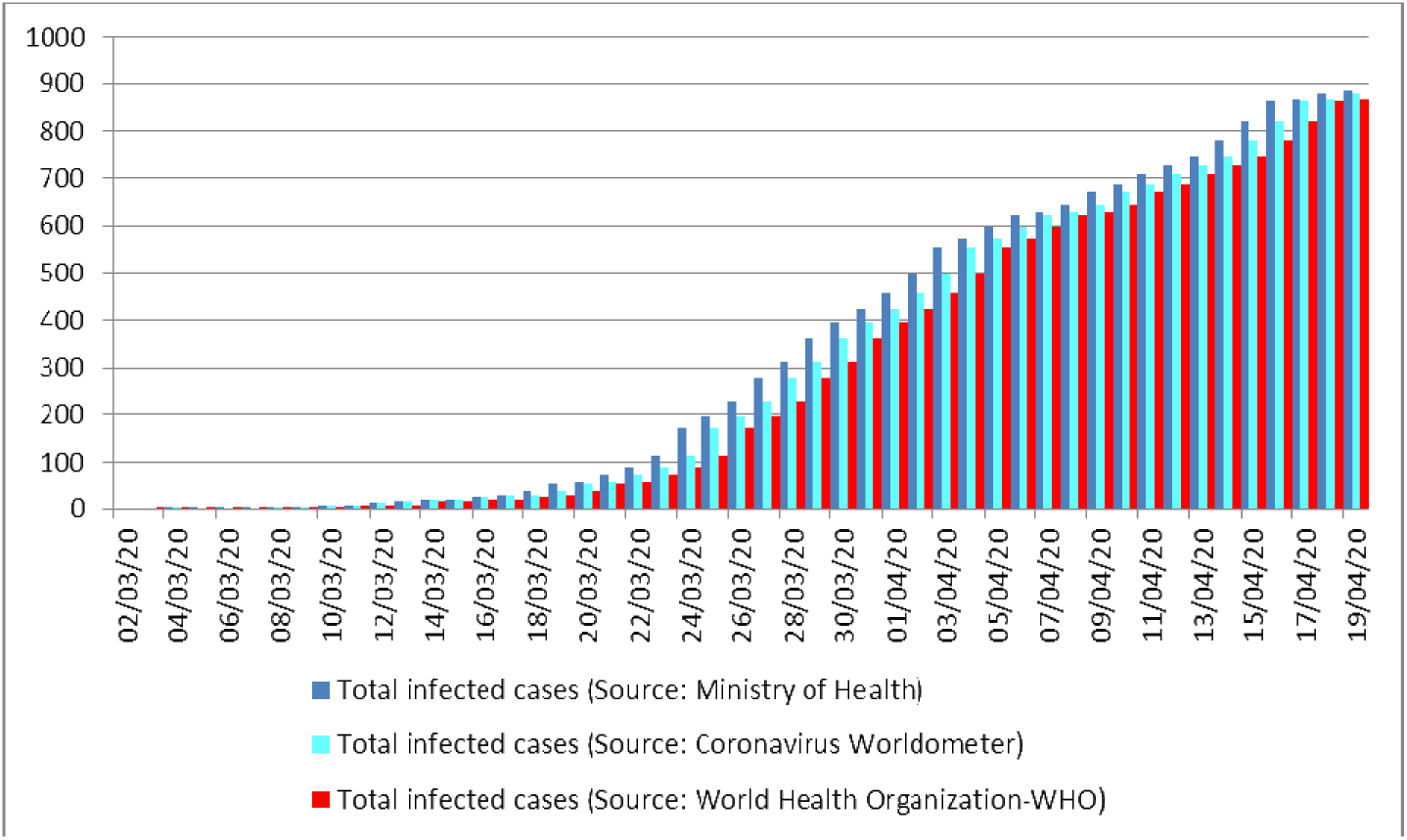
Times series of the total infected cases reported from March 02, 2020 to April 19, 2020.

**Figure 2.**
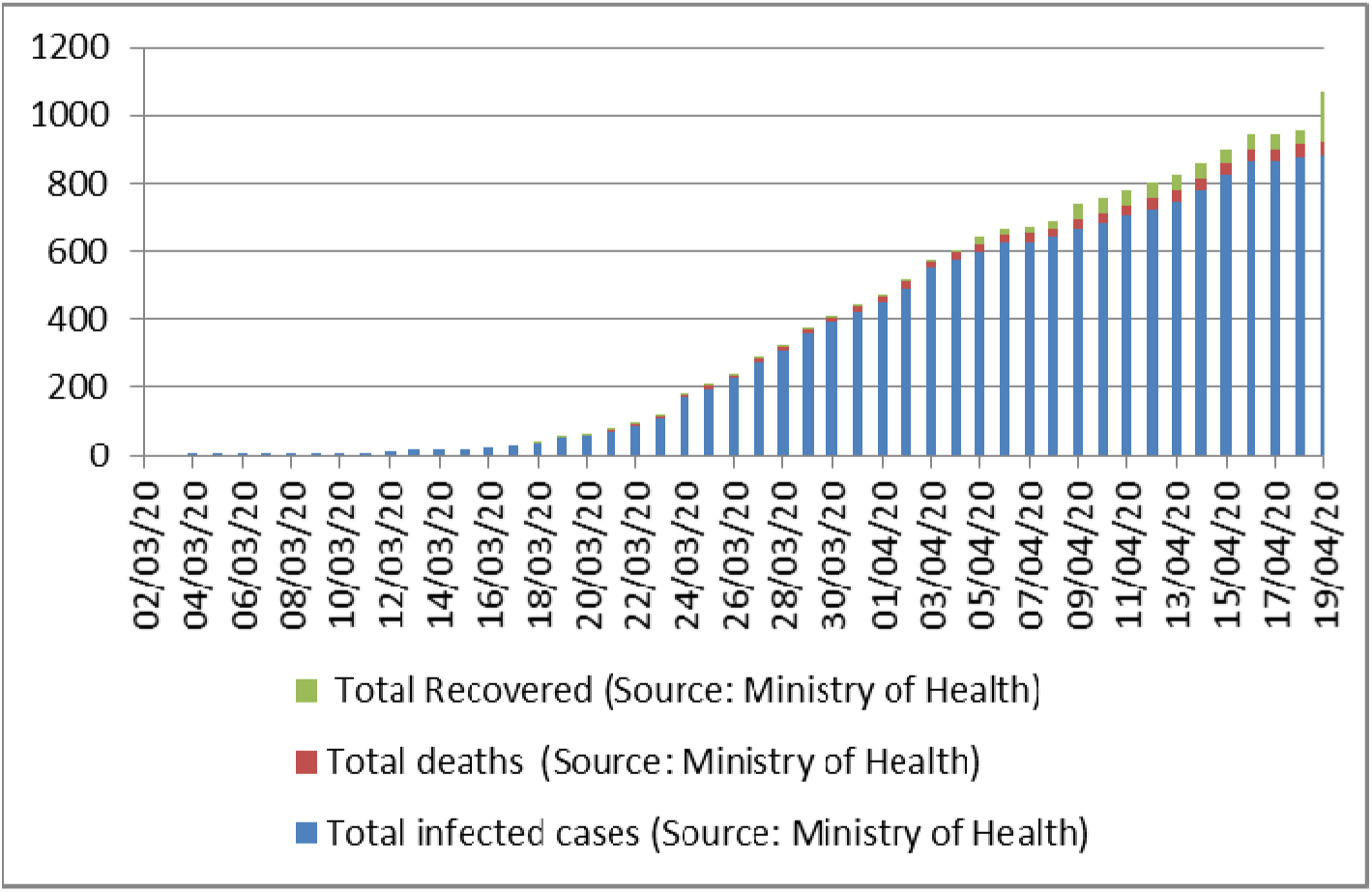
Times series of the total infected cases, total deaths, and total recovery 02, 2020 to April 19, 2020.

**Figure 3.**
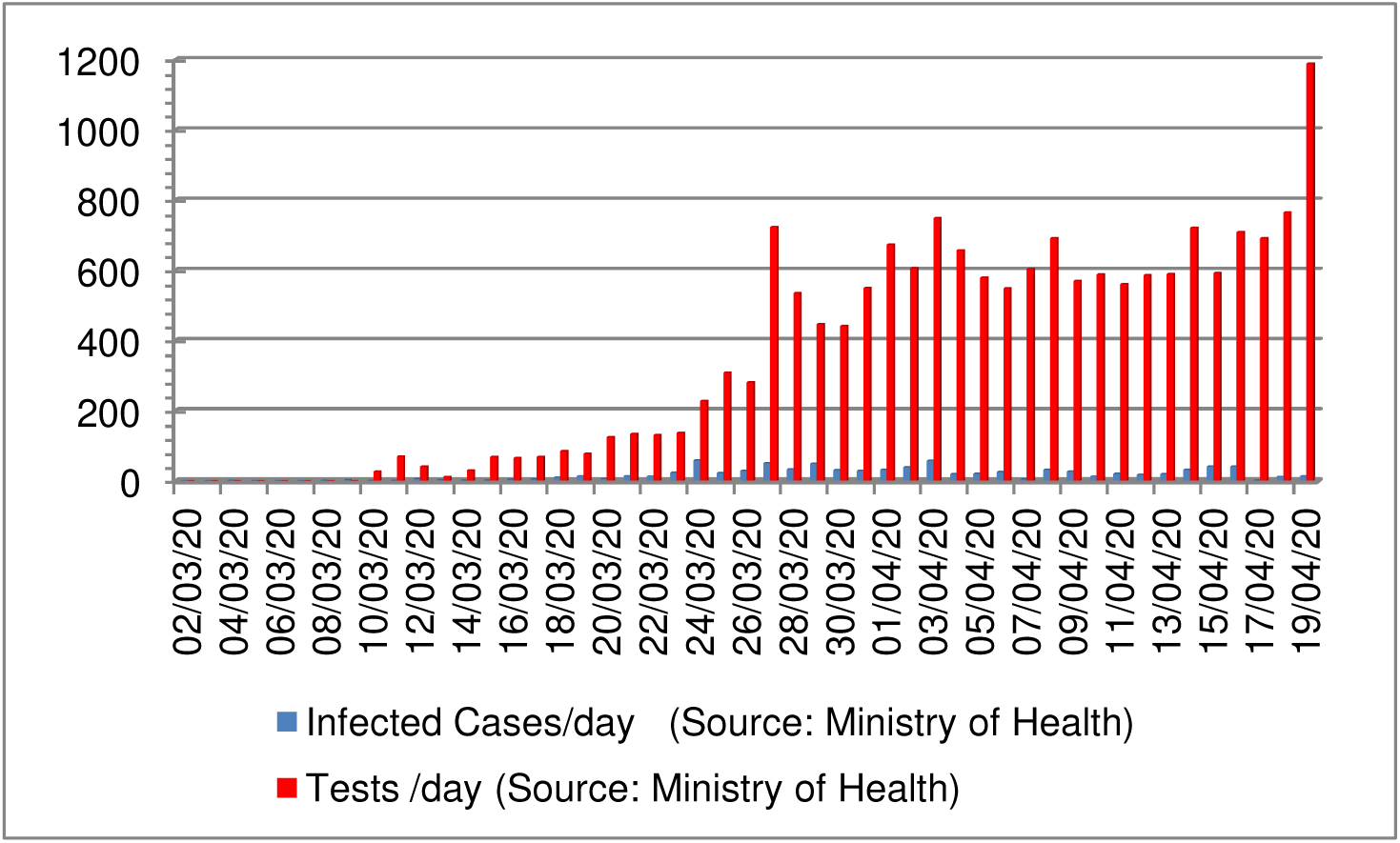
Time series of the daily tests and the infected cases/day reported from March 02, 2020 to April 19, 2020.

The SIR model was then compiled using the dataset [1] consisting of 45 number determinations. Fig. 4 shows the actual and predicted total number of cases using the SIR model in Tunisia. Fig. 5 shows the actual and predicted daily number of cases. Although the estimated decline begins on April 14, the curve shows a tail until May 10, 2020 for zero new infection. The estimated basic reproduction number 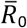 is equal to 2.025, where the epidemic peak has occurred on or around first April 2020.

**Figure 4.**
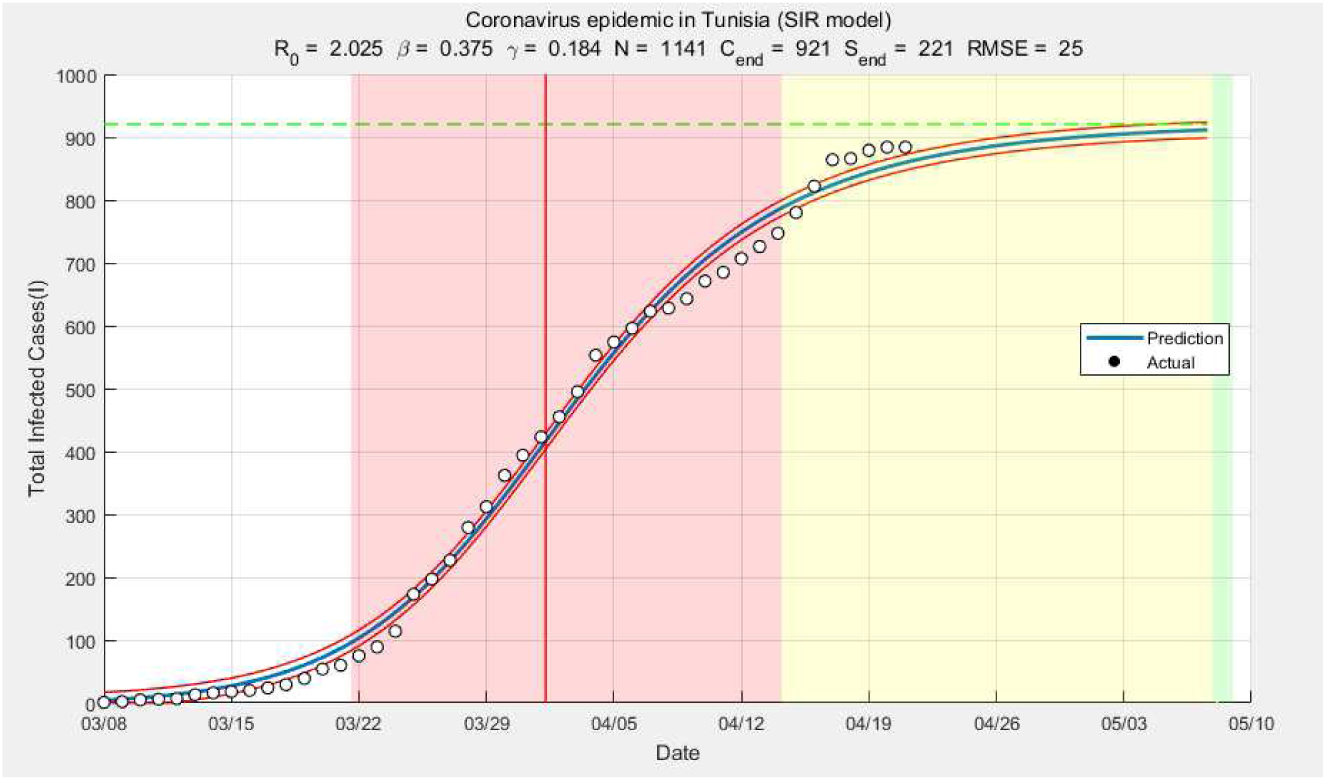
Actual and predicted total number of cases using the SIR model in Tunisia. Cend represents the epidemic size, and Send represents the number of susceptible individuals left. RMSE is the root mean square error.

**Figure 5.**
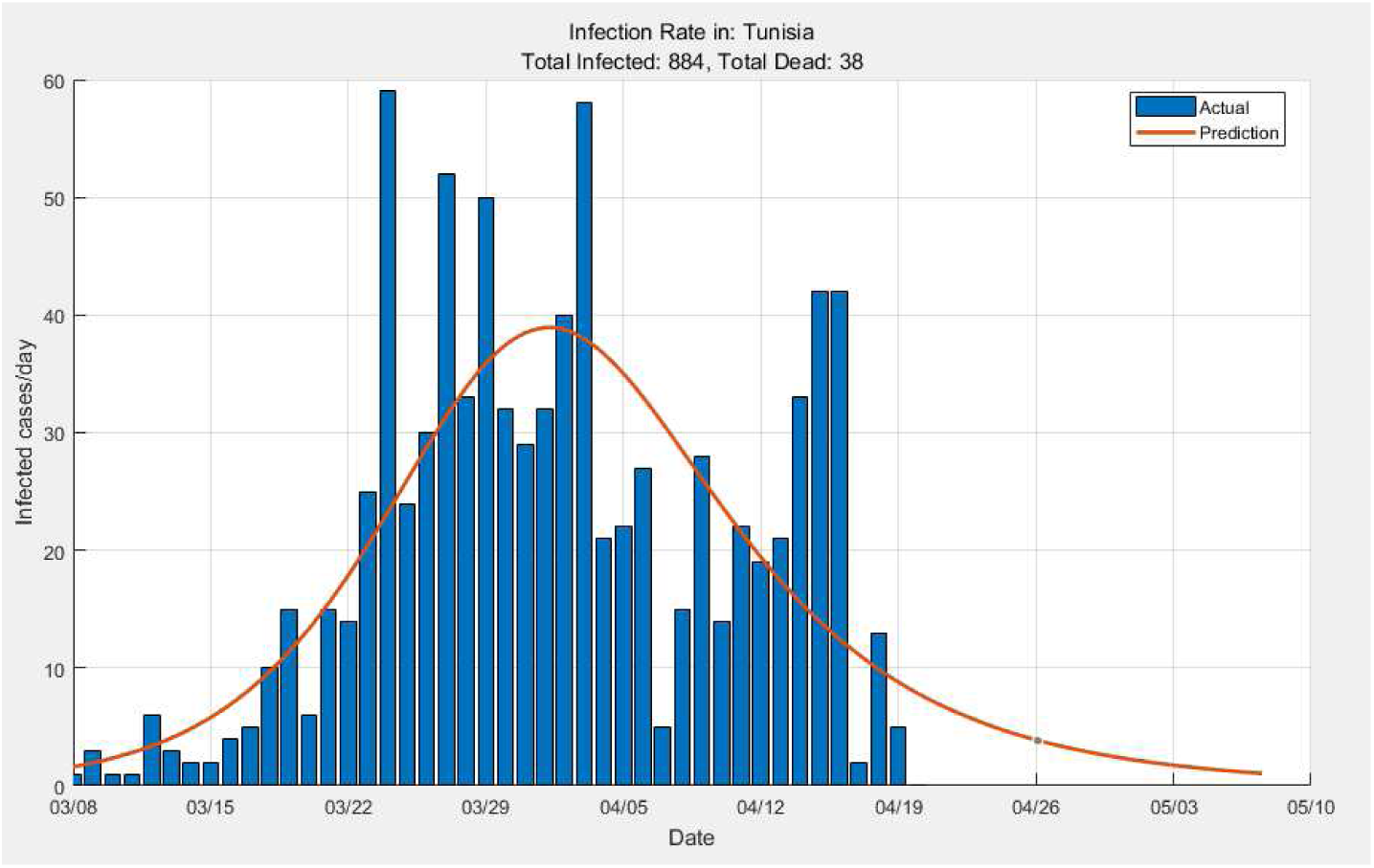
Actual and predicted daily number of cases using the SIR model in Tunisia.

The obtained results are very useful in this stage for two reasons: 1) validate the rigorous and wise strategy undertaken by the Tunisian authorities versus real facts described in Fig. 6, and 2) verify if anti-lockdown scheduled for May 04, 2020 is well advised. So far, it seems that the results of the containment policy adopted by the national authorities are very satisfactory compared to the situations currently experienced by countries neighboring the Magreb and the Middle East reported in [2,3]. We hope that this situation will be stable and durable and will not be destabilized by anti-lockdown.

**Figure 6.**
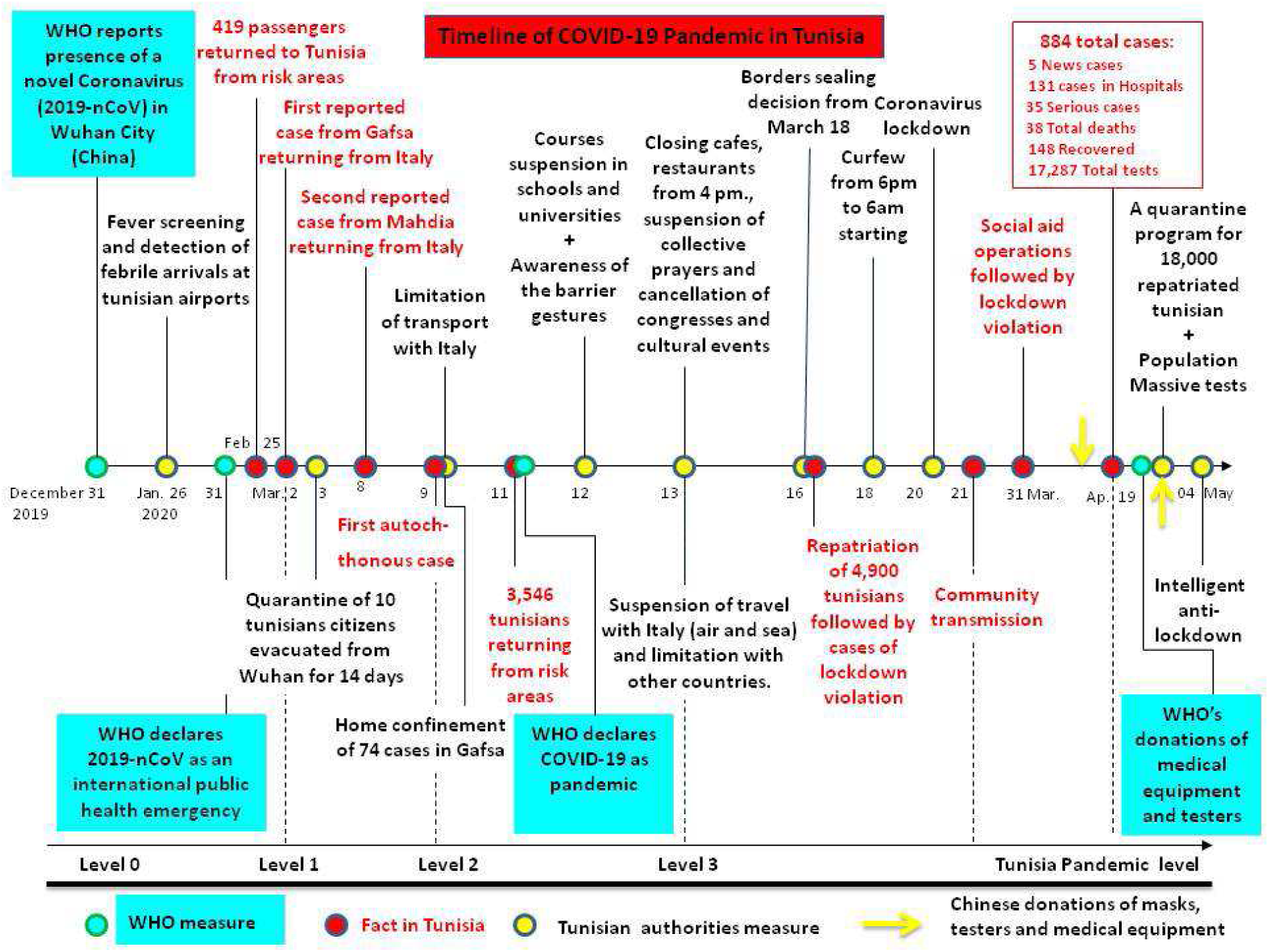
Timeline of the Covid-19 in Tunisia: A useful tool for mathematical modeling and forecasting [6].

## Conclusion and future work

In this paper a dataset is collected for the COVID-19 epidemic situation Tunisia from March 02, 2020 to April 19, 2020, and then applied to the SIR model. The obtained results estimated and predicted the dynamics of the epidemic spread and evaluated the authorities adopted strategies. The designed model consolidates the successful Tunisian experience in fighting the outbreak. In future work, a more complex reliable model will be designed taking on account of the timeline of the Covid-19 in Tunisia in order to help control the spread.

## Data Availability

Data can be retrieved from the public repository:
Mendeley Data
Data identification number: 10.17632/v8fhn6wsh2.3
Direct URL to data: https://data.mendeley.com/datasets/v8fhn6wsh2/3

https://doi.org/10.17632/v8fhn6wsh2.3

## Acknowledgments

This work was partially supported by the National Institute of Applied Sciences and Technology of Tunis, Tunisia.

